# Diminished responses to mRNA-based SARS-CoV-2 vaccines in individuals with rheumatoid arthritis on immune modifying therapies

**DOI:** 10.1101/2023.01.03.23284167

**Authors:** Samuel D. Klebanoff, Lauren B. Rodda, Chihiro Morishima, Mark H. Wener, Yevgeniy Yuzefpolskiy, Estelle Bettelli, Jane H. Buckner, Cate Speake, Marion Pepper, Daniel J. Campbell

## Abstract

Rheumatoid arthritis (RA) is a chronic inflammatory autoimmune disorder that causes debilitating swelling and destruction of the joints. People with RA are treated with drugs that actively suppress one or more parts of their immune system, and these may alter their response to vaccination against SARS-CoV-2. In this study, we analyzed blood samples from a cohort of RA subjects after receiving a 2-dose mRNA COVID-19 vaccine regimen. Our data show that individuals on the CTLA4-Ig therapy abatacept have reduced levels of SARS-CoV-2-neutralizing antibodies after vaccination. At a cellular level, these subjects show reduced activation and class-switching of SARS-CoV-2-specific B cells, as well as reduced numbers and impaired helper cytokine production by SARS-CoV-2-specific CD4^+^ T cells. Individuals on methotrexate showed similar but less severe defects in vaccine response, whereas individuals on the B cell-depleting therapy rituximab had a near-total loss of antibody production after vaccination. These data define a specific cellular phenotype associated with impaired response to SARS-CoV-2 vaccination in RA subjects on different immune-modifying therapies, and help inform efforts to improve vaccination strategies in this vulnerable population.

## Introduction

The COVID-19 pandemic, caused by Severe Acute Respiratory Syndrome Coronavirus 2 (SARS-CoV-2), has resulted in over 6 million deaths and worldwide economic and social disruption. Vaccines targeting the SARS-CoV-2 spike (S) protein are essential tools in combating this pandemic, and have proved highly efficacious in preventing severe disease, hospitalization, and death. In the United States, over two thirds of the population has been vaccinated, with the two most common vaccines being Pfizer’s BNT162b2 and Moderna’s mRNA-1273 vaccines which use modified mRNA platforms that induce potent cellular and humoral responses to the S protein (1, 2). However, for patients with a compromised immune system, such as those with autoimmune disease taking immunosuppressive therapies, vaccination can often be less effective (3). Although both vaccines showed ∼95% efficacy at preventing COVID-19 in initial clinical trials, immunocompromised patients were excluded from those trials (4), and a better understanding of the response to COVID-19 vaccination in this patient population is urgently needed. This is especially true given the emergence of viral variants that partially evade antibody-mediated protective immunity due to structural mutations in the S protein.

The response to SARS-CoV-2 mRNA vaccines is characterized by rapid production of S protein-specific antibodies, initially from short-lived plasmablasts and later from a smaller pool of long-lived plasma cells (5, 6). The majority of vaccine-induced neutralizing antibodies target the S protein receptor binding domain (RBD) and contribute to protection by preventing interaction with the ACE2 receptor on human epithelial cells, thus blocking infection. Serum levels of anti-S antibodies decline slowly over the course of several months but rebound quickly upon administration of subsequent booster vaccine doses or reinfection as S-specific memory B cells generated by the initial vaccination rapidly activate and differentiate into antibody-secreting plasmablasts (5). Vaccination also induces strong CD4^+^ and CD8^+^ T cell responses, as measured by expression of activation markers such as CD69 and CD137 by these cells after stimulation with S protein peptides. Among CD4^+^ T cells, effector and memory T cells producing key anti-viral cytokines such as IL-2, IFNγ and IL-21 dominate the response, and an expanded population of S-specific T cells persists for at least several months after vaccination (5, 7).

Patients with autoimmune diseases such as rheumatoid arthritis (RA) are treated with drugs that target key immune pathways important for disease pathology, but that can also impair effective vaccine responses. Indeed, although the American College of Rheumatology has recognized the potential of these therapies to impact SARS-CoV-2 vaccination, there is limited consensus on whether to recommend brief cessation of treatment for RA patients receiving the SARS-CoV-2 vaccines (8). Conventional disease-modifying anti-rheumatic drugs (DMARDs) are anti-inflammatory and immunosuppressive small molecule drugs, the most common of which is methotrexate (MTX) which has become the standard-of-care for RA. The mechanism of action of MTX in RA has not been fully defined, although it is thought to act via adenosine signaling and blocking folate metabolism in disease-causing lymphocytes (9, 10). Patients whose disease is difficult to control with MTX and other first-line treatments are also treated with recombinant biologic drugs, among which is the CTLA4-Ig therapy abatacept. Abatacept functions by binding to CD80 and CD86 on antigen-presenting cells, effectively blocking their ability to provide co-stimulation to pathogenic autoreactive T cells. We and others demonstrated that abatacept treatment reduces the number and activity of circulating T follicular helper (Tfh) T cells (11-13), a specialized CD4^+^ T cell population that produces IL-21 and provides help to promote the proliferation, isotype class switching and affinity maturation of antigen-specific B cells (14). Indeed, co-stimulation blockade via abatacept inhibits vaccine-induced antibody responses, including to SARS-CoV-2 mRNA vaccines (15-17). However, a detailed cellular analysis of T and B cell responses to SARS-CoV-2 vaccination in RA patients on these therapies is still lacking. Rituximab, an anti-CD20 antibody which depletes B cells, is also used to treat RA, and as expected, individuals treated with rituximab have severely blunted vaccine responses, including to SARS-CoV-2 mRNA vaccines (18, 19).

For this study, we assembled a cohort of individuals with RA who were treated with MTX, abatacept, or rituximab and compared their responses to SARS-CoV-2 vaccination with healthy control subjects. We measured S protein-specific antibody responses in the serum, and assessed the abundance, phenotype and function of SARS-CoV-2-specific T cells and B cells. We found that all cohorts of individuals with RA had altered vaccine responses compared to healthy controls. As expected, the lack of B cells resulted in a near-total loss of anti-SARS-CoV-2 antibodies in rituximab-treated individuals. Abatacept treatment also led to reduced S-specific and neutralizing antibodies. Interestingly, the number of RBD-specific B cells found in peripheral blood was similar in control, abatacept- and MTX-treated subjects. However, abatacept reduced B cell class switching to IgG and altered memory B cell differentiation. The number of SARS-CoV-2-specific CD4^+^ T cells was decreased in MTX- and abatacept-treated subjects, and production of the key cytokines IL-2, IFNγ and IL-21 was also diminished by abatacept treatment. Thus, abatacept treatment limits the efficacy of SARS-CoV-2 mRNA vaccines in a manner consistent with impaired generation of optimal T cell responses capable of providing help to B cells for production of high-titer, class-switched virus-neutralizing antibodies. Understanding the mechanistic basis for these impaired responses sheds light on the cellular networks required for immune protection in SARS-CoV-2 vaccinated individuals. These results also provide additional support for temporary cessation of abatacept treatment before vaccination when clinically manageable to help ensure optimal vaccine-induced immune protection from SARS-CoV-2 infection.

## Results

### Abatacept and rituximab reduce humoral immune responses to SARS-CoV-2 vaccination

Our study cohort consisted of 40 individuals, including 13 healthy controls and 27 subjects with rheumatoid arthritis (RA) (Fig. 1A). 11 participants with RA were being treated with MTX, 11 were being treated with abatacept (6 of whom were also on additional therapies including hydroxychloroquine, prednisone, leflunomide, and sulfasalazine), and 5 were being treated with rituximab (3 of whom were also on additional therapies including MTX, hydroxychloroquine, and leflunomide). Healthy controls were selected to be approximately age- and sex-matched to the RA cohort. All study participants donated a single blood sample after receiving the second dose of either the Pfizer BNT162b2 or Moderna mRNA-1273 vaccines. Donors were requested to provide a blood sample within 1-3 weeks after vaccination, but in some cases participants with RA donated blood at their next clinical visit – which generally occurred within 3 months of vaccination. For all blood samples collected, serum and PBMCs were isolated and subjected to humoral and cellular analyses (Fig. 1A).

**Figure 1.**
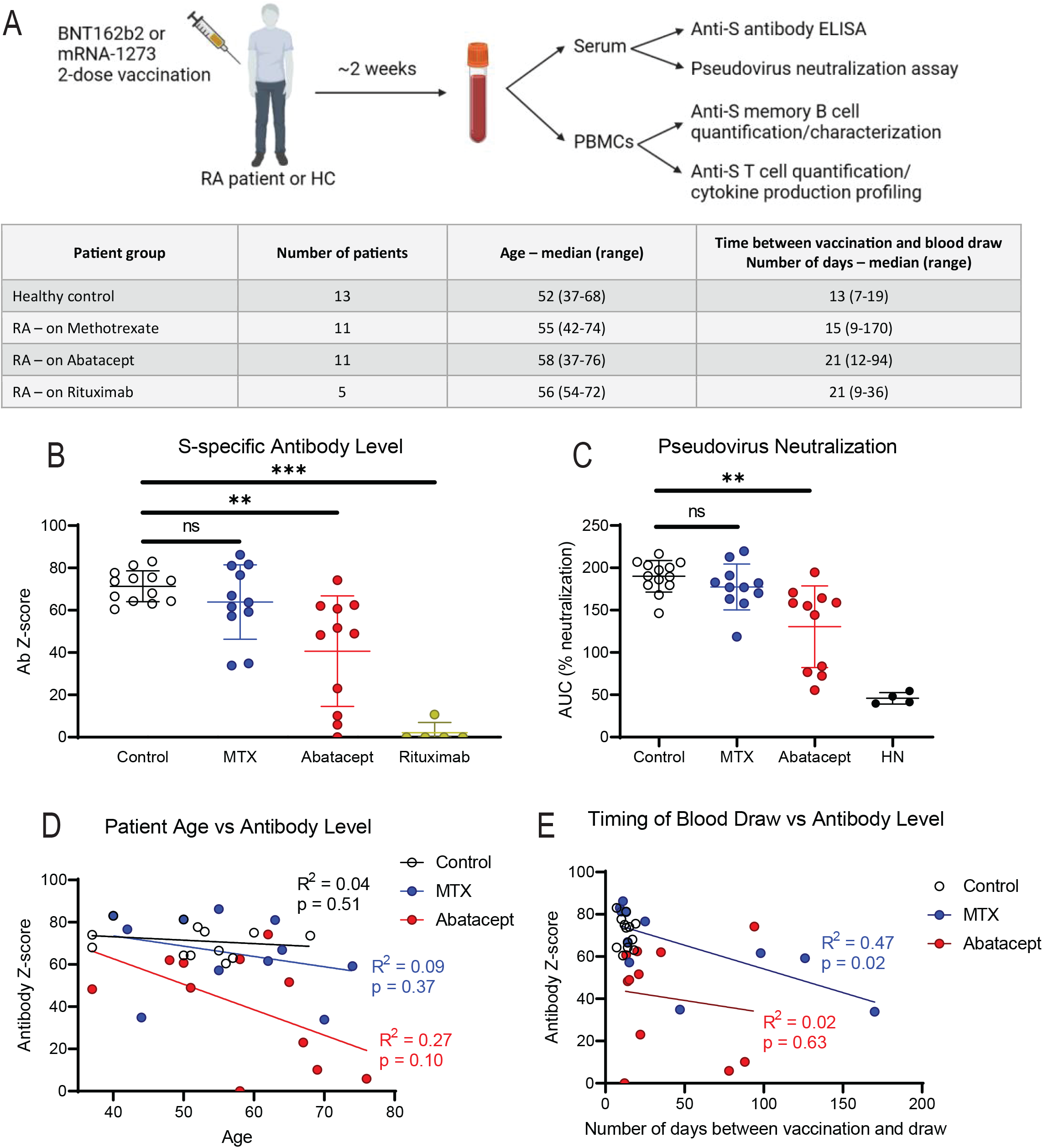
SARS-CoV-2-specific Antibodies and Pseudovirus Neutralization are Reduced in RA Patients After Vaccination. **(A)** Study schematic and table of participant information. **(B)** Normalized anti-S antibody levels as measured by ELISA. **(C)** Pseudovirus neutralization of subjects’ sera, as area-under-curve (AUC) across serum dilutions, with historical/naive (HN) control. **(D)** Subject age graphed against anti-S antibody levels. **(E)** Time between each subject’s second vaccine dose and blood draw for the study graphed against anti-S antibody levels. *Linear regression shown with r-squared values and p values testing probability of a non-zero slope. Statistics determined by Kruskal-Wallis test with post-hoc Dunn’s multiple comparison test. * p<*.*05, ** p<*.*01, *** p<*.*001*

Generation of virus-neutralizing antibodies is the primary goal of vaccination and these correlate strongly with protection from SARS-CoV-2 (6, 20). Using ELISA to measure S-protein specific IgG and normalizing to a historical negative control group (Fig. 1B), we found as expected that patients on the B cell-depleting therapy rituximab showed almost no detectable level of antibodies in their serum, whereas patients on abatacept generated significantly lower levels of S-specific antibodies than healthy controls subjects. We did not observe a significant decrease in the antibody response in MTX-treated subjects, although responses trended lower in a subset of these individuals.

In addition to measuring anti-S antibody levels, we conducted a pseudovirus neutralization (pVNT) assay in which lentiviral particles pseudotyped with the SARS-CoV-2 S protein are incubated with serum to measure blockade of infection of ACE2-expressing target cells. Samples from SARS-CoV-2 naïve and unvaccinated individuals drawn in early 2020 were included as negative controls, as well as a monoclonal anti-RBD antibody as a positive control (Fig S1A). As with the total antibody levels, we found that abatacept, but not MTX, significantly decreased serum neutralization activity compared with healthy control subjects (Fig. 1C). Indeed, we observed a strong correlation between total anti-S antibody IgG levels and pseudovirus neutralization among our subjects (Fig S1B), indicating that although quantitatively impaired, the quality of antibody produced in MTX- and abatacept-treated subjects was largely normal.

Immune function declines with age, and therefore patient age is a potentially confounding variable in our study. In healthy control and MTX-treated subjects, anti-S antibody levels showed no discernable correlation with patient age, whereas in abatacept-treated subjects there was a slight negative correlation with age that was not statistically significant (Fig. 1D). Another potentially confounding variable is the time between completion of the vaccine series and sample acquisition for our study. This variable is particularly important to address since our control samples were all obtained within 3 weeks of vaccination, whereas samples from individuals with RA were collected as late as 6 months post-vaccination. However, we did not observe a significant correlation between the time of sample collection and anti-S antibody levels or pseudovirus neutralization activity in either the MTX- or abatacept-treated groups (Fig 1E, Fig S1C). Thus, differences in age or sample timing do not account for the diminished antibody production we observed in abatacept-treated subjects. Additionally, we found no difference in antibody production between subjects on abatacept mono-therapy vs subjects on abatacept in combination with any other therapy (Fig S1D).

### Abatacept reduces activation and class-switching of RBD-specific memory B cells in response to SARS-CoV-2 vaccination

Diminished antibody responses to SARS-CoV-2 vaccination indicate that the activation and functional differentiation of SARS-CoV-2-specific B cells in abatacept-treated subjects may be altered. Therefore, we used RBD tetramer probes to characterize vaccine-induced RBD-specific B cell responses in blood samples from our MTX- and abatacept-treated RA cohorts (20). We used a decoy tetramer (containing all elements of the tetramer except the RBD) to control for non-specific binding, and performed magnetic enrichment to increase the frequency of RBD-binding among analyzed cells (Fig 2A, left). Despite diminished antibody responses in subjects on abatacept, the total numbers of RBD-specific B cells were statistically similar among all our cohorts (Fig. 2A, right, Fig S2A).

**Figure 2.**
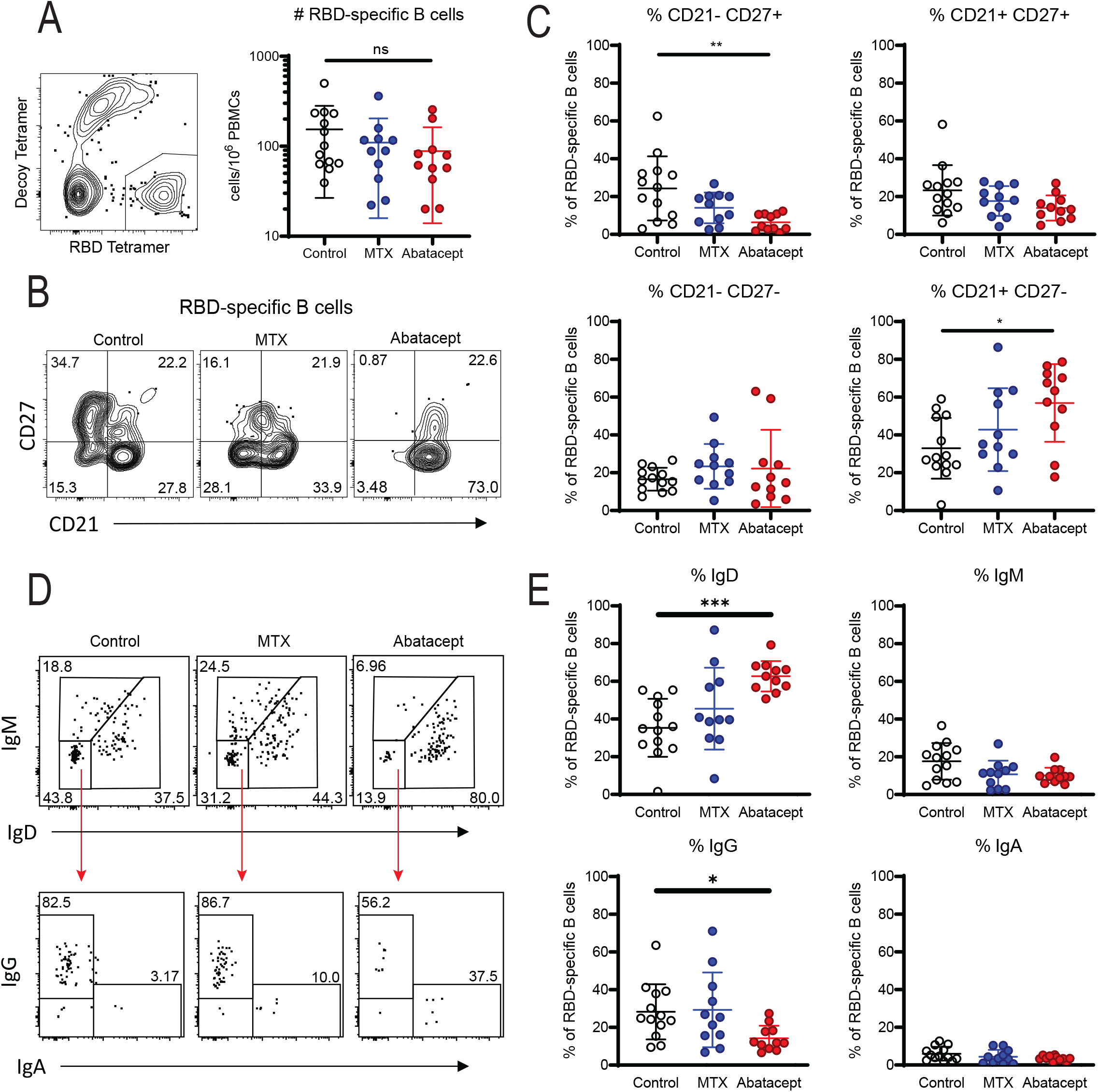
RBD-specific Memory B Cell Response is Weaker in Abatacept Patients After Vaccination. **(A)** Representative gating on live CD3-CD14-CD16-CD19+CD20+ B cells (left) and number (right) of SARS-CoV-2 RBD specific B cells (RBD tetramer+ Decoy tetramer−) from PBMCs from control (white), methotrexate (MTX, blue) and abatacept (red) treated individuals. **(B)** Representative gating on RBD-specific CD38lo non-plasmablast B cells for naïve B cells (CD21+CD27−), classical MBCs (CD21+CD27+), activated MBCs (CD21−CD27+) and double-negative (DN) activated MBCs (CD21−CD27−). **(C)** Proportion of RBD-specific B cells that are each phenotype from individuals in the indicated treatment group. **(D)** Representative gating on RBD-specific CD38lo non-plasmablast B cells for isotypes IgD, IgM, IgG and IgA. **(E)** Proportion of RBD-specific B cells expressing the isotypes indicated in the groups indicated. Data combined from four individual experiments. *Statistics determined by Kruskal-Wallis test with post-hoc Dunn’s multiple comparison test. All statistically significant comparisons (p<*.*05) are shown. * p<*.*05, ** p<*.*01, *** p<*.*001*

After detecting their antigen, CD21^+^CD27^-^ naïve B cells proliferate and become CD21^-^CD27^+/-^ activated B cells and then differentiate into resting CD21^+^CD27^+^ classical memory B cells (MBCs) which can rapidly produce protective antibodies upon a reinfection (21). Following vaccination, abatacept treatment was associated with a significantly lower proportion of CD21^-^ CD27^+^ RBD-specific B cells and a higher proportion that retained a CD21^+^CD27^-^ naïve phenotype compared to healthy control or MTX cohorts (Fig. 2B and 2C). Since the proportion of activated memory B cells declines with time after vaccination (5), we tested the contribution of timing to the depressed RBD-specific B cell activation in the abatacept group by correlating this proportion to the time of blood draw post-vaccination. While these factors were negatively correlated for healthy controls (Fig. S2B), there was no correlation with time-post vaccination in either the MTX or abatacept cohorts (Fig S2C), suggesting timing does not explain the difference in activation. In contrast, the proportion of classical MBCs (CD21^+^CD27^+^) was not different between controls and treated patients (Fig. 2C). Finally, vaccination did not induce significant proportions of CD21^-^CD27^-^CD11c^+^ atypical memory B cells, which are associated with aberrant B cell activation in some viral infections (22), in any of our subject groups (Fig. S2D). Thus, the phenotypic changes we observe are reflected in fewer antigen-experienced activated and memory RBD-specific B cells generated by vaccination in the context of abatacept treatment (Fig. S2E), although there was not a significant correlation between the number of antigen-experienced MBCs and level of anti-S antibodies in serum from individuals on MTX or abatacept (Fig S2F).

The effector function of antibodies depends on their isotype, and the generation of virus-neutralizing IgG is a primary correlate of disease protection in the context of SARS-CoV-2 vaccination (23, 24). Abatacept can interrupt the differentiation and function of Tfh that help drive BCR class switching from IgD and IgM to predominantly IgG and IgA. Therefore, we assessed the isotype of RBD-specific B cells in each subject group (Fig 2E). While the MTX cohort showed no significant differences in antibody class switching among RBD-specific B cells compared with controls, abatacept treatment was associated with a significantly lower percentage of IgG^+^ cells, and a higher percentage of IgD^+^ cells. This is consistent with the reduced B cell activation and increased proportion of naïve-phenotype cells in these subjects. However, even within the antigen-experienced B cell population, we found abatacept-treated subjects had a significantly higher percentage of unswitched IgD^+^ cells and a trend towards a lower percentage of IgG^+^ cells (Fig S2G). This indicates that abatacept treatment impairs signals that lead to class-switching in addition to those that support naïve B cell activation and differentiation into memory.

### MTX and abatacept impair development of S-specific memory T cells after SARS-CoV-2 vaccination

To determine whether the CD4^+^ T cell response to SARS-CoV-2 vaccination in individuals with RA was impaired compared to controls, we stimulated PBMCs overnight with a pool of peptides from the SARS-CoV-2 S protein, and then stained the cells with a T cell activation-induced marker (AIM) flow cytometry panel. We included a negative control vehicle-only stimulation condition (DMSO), and used the commercially available CEFX peptide pool that contains 68 known peptide epitopes from 18 common pathogens that reliably stimulates T cells across a broad range of HLA haplotypes as a positive control (25). We also stimulated cells with a peptide pool from the SARS-CoV-2 membrane/nucleocapsid (M/N) proteins that induces a robust response in individuals previously infected with SARS-CoV-2, but not in vaccinated subjects (5).

We used CD69 and CD137 as representative AIMs, the co-expression of which indicated that a T cell had become activated and was therefore specific for one of the peptides in the stimulation condition (Fig 3A). All groups showed similar CD4^+^ T cell responses to the CEFX pool, and there was no detectible responses to the M/N pool (Fig S3A), confirming that the subjects in our cohorts were not previously infected with SARS-CoV-2. However, the frequency of vaccine-induced S-specific activated T cells was significantly reduced in MTX-treated subjects, and also trended lower in the abatacept cohort (Fig 3B). Rituximab-treated subjects were also included in these analyses and had lower levels of S-specific AIM^+^ T cells compared to controls (Fig S3B). However, due to the small sample size and low numbers of activated cells, this group was excluded from subsequent phenotypic analyses.

**Figure 3.**
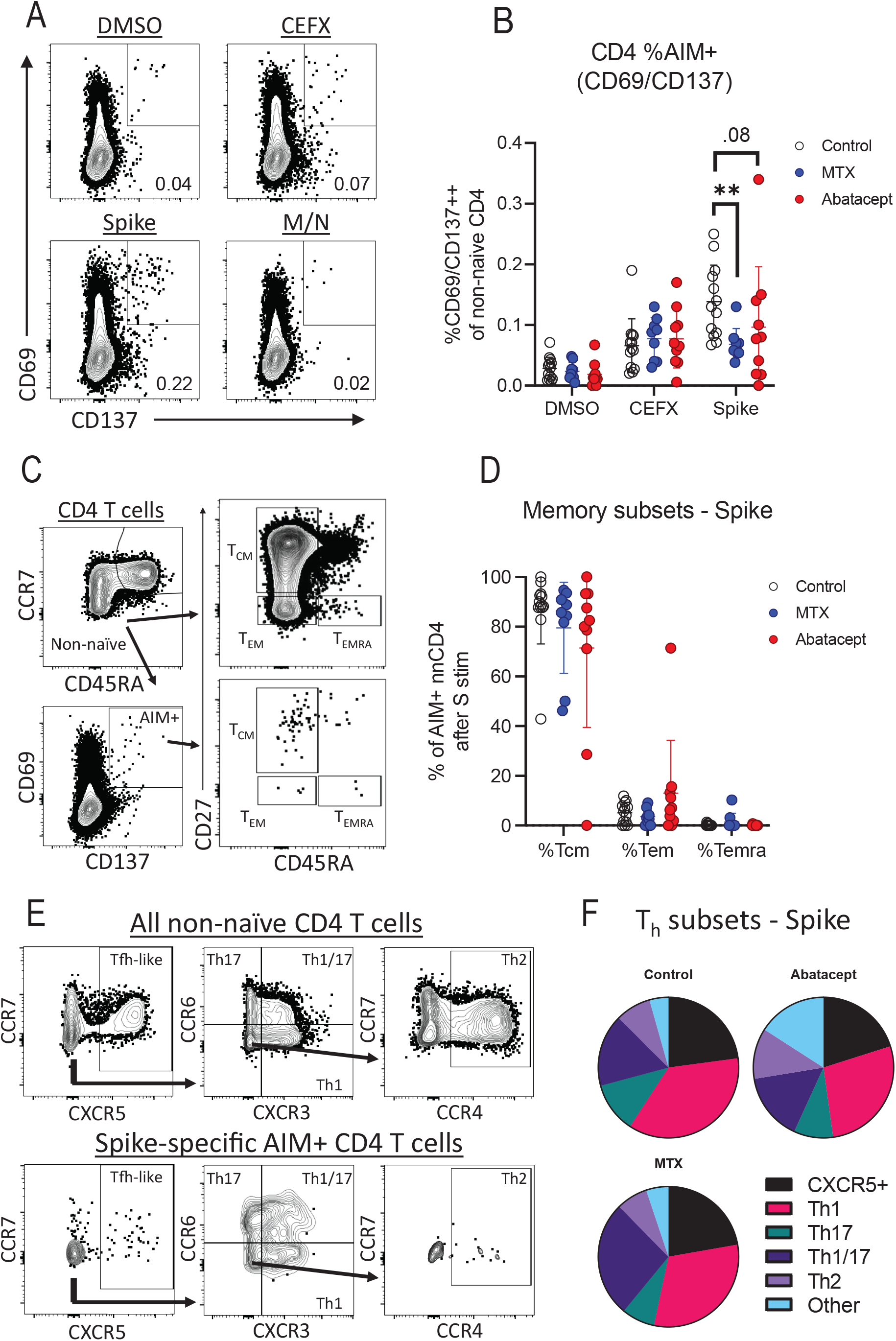
RA Patients have Reduced Numbers but Similar Phenotype of Spike-specific T Cells. **(A)** Representative gating of CD3+CD45RA-CD4+ T cells for AIM+ (CD69+CD137+) within indicated stimulation conditions. **(B)** Quantification of AIM expression by subject groups as percentage of CD3+CD45RA-CD4+ cells. **(C)** Representative gating of central memory (CD45RA-CD27+), effector memory (CD45RA-CD27-), and Temra (CD45RA+CD27+) within non-naïve and AIM+ T cells. **(D)** Quantification of CD4 memory subsets within spike-stimulated AIM+ cells. **(E)** Representative gating of CXCR5+ (containing the Tfh population), Th1 (CXCR3+CCR6-), Th17 (CXCR3-CCR6+), Th1/17 (CXCR3+CCR6+), and Th2 (CXCR3-CCR6-CCR4+) cells. **(F)** Pie charts showing percentage of spike-stimulated AIM+ CD4 T cells falling into each Th subset. *Statistics determined by Kruskal-Wallis test with post-hoc Dunn’s multiple comparison test. All statistically significant comparisons (p<*.*05) between treatment groups are shown. * p<*.*05, ** p<*.*01, *** p<*.*001*

We next performed phenotypic characterization of the S-specific T cells, first breaking down the non-naïve CD4^+^ T cells into central memory (CD27^+^CD45RA^-^), effector memory (CD27^-^ CD45RA^-^) and T_EMRA_ (CD27^-^CD45RA^+^) subsets (Fig. 3C). We observed no significant differences in the proportion of these memory populations in MTX- or abatacept-treated RA participants compared with healthy controls (Fig. 3D). We also used differential chemokine receptor expression to identify which functionally CD4 T helper subsets were represented in the AIM^+^ cells as previously described (5). In this analysis, we also found no significant difference between groups in the percentage of S-specific CD4^+^ T cells falling into any functional Th subset, and in all cohorts CXCR3^+^ Th1 cells and CXCR3^+^CCR6^+^ Th1/17 cells dominated the response (Fig. 3E,F). However, the phenotypic breakdown of S-specific T cells in individual subjects varied widely, and the small numbers of AIM^+^ T cells in many patients reduced our ability to find significant differences between groups (Fig. S4).

### Reduced production of Tfh-associated cytokines by S-specific T cells from abatacept-treated subjects

In addition to the phenotype of S-specific CD4^+^ T cells, we interrogated the ability of these cells to produce anti-viral cytokines upon restimulation by performing intracellular cytokine staining on PBMCs stimulated with the S peptide pool. For these experiments, we used CD69 and CD154 as activation markers to identify S protein-specific T cells (Fig 4A), and CD69^+^CD154^+^ activated cells are the primary cytokine producing cells when restimulated with spike peptide (5, 20). Although the timing of peptide stimulation is much shorter than in our AIM assay (six hours vs overnight), and CD154 is used to identify activated cells rather than CD137, we found that the relative numbers of activated CD4^+^ T cells in this assay followed the same trend as in our AIM assay using CD69 and CD137 to identify S-specific cells (Fig S5).

**Figure 4.**
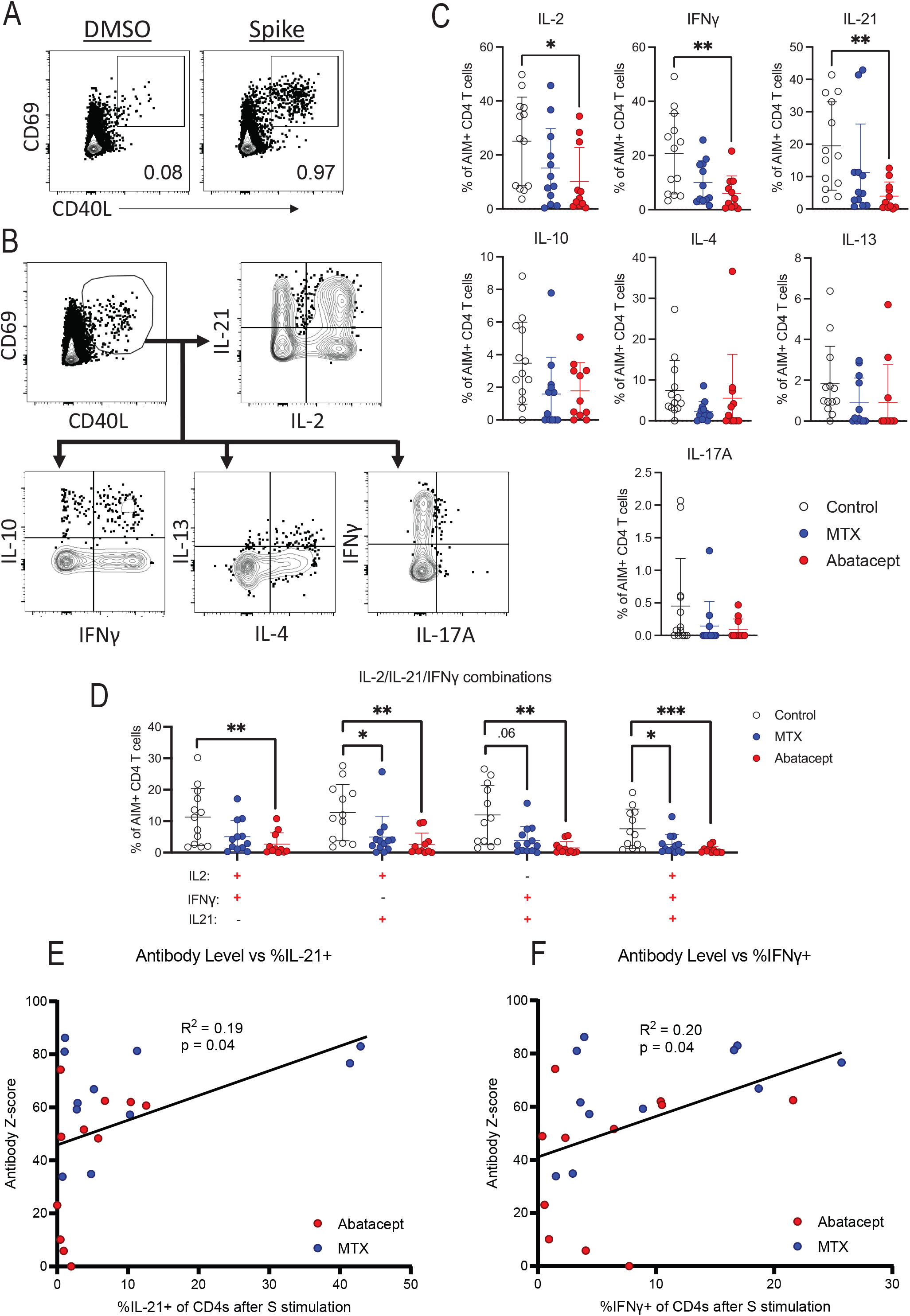
Intracellular Cytokine Staining of Patient T Cells After Brief Spike Peptide Stimulation. **(A)** Representative gating of AIM+ (CD69+CD40L+) T cells for ICS assay co-culture. **(B)** Representative gating of IL-2, IL-21, IL-10, IFNγ, IL-4, IL-13, and IL-17A expression within AIM+ CD4 T cells. **(C)** Quantification of the expression of each cytokine by percentage of AIM+ CD4s. **(D)** Co-expression of IL-2, IL-21, and IFNγ in each indicated combination. **(E)** Anti-S antibody level graphed percent of spike-activated AIM+ CD4s expressing IL-21. **(F)** Anti-S antibody level graphed percent of spike-activated AIM+ CD4s expressing IFNγ. *Linear regression shown with r-squared values and p values testing probability of a non-zero slope. Statistics determined by Kruskal-Wallis test with post-hoc Dunn’s multiple comparison test. All statistically significant comparisons (p<*.*05) are shown. * p<*.*05, ** p<*.*01, *** p<*.*001*

We analyzed the expression of the cytokines IL-2, IFNγ, IL-4, IL-13, IL-17A, IL-21, and IL-10 by S-specific CD4 memory T cells from each subject (Fig. 4B). As previously reported the cytokine response to SARS-CoV-2 was heterogenous (5), and we observed substantial fractions of S-specific cells producing each of the analyzed cytokines other than IL-17A. Moreover, we found that the fractions of cells producing IL-2, IFNγ, or IL-21 were significantly reduced in abatacept-treated subjects compared to healthy controls (Fig. 4C), and especially the proportion of cells co-expressing IL-21, IL-2 and IFNγ was highly reduced in both MTX- and abatacept-treated participants (Fig 4D). IL-21 production is critical for Tfh cell function, and IFNγ promotes class switching to IgG, suggesting that these T cell defects may be linked to the relatively poor B cell and antibody responses we observed in MTX- and abatacept-treated subjects. Indeed, expression of either IL-21 or IFNγ correlated with S-specific antibody levels in serum of MTX- and abatacept-treated subjects (Fig 4E, F), indicating that these therapies impair humoral immunity by disrupting T cell-B cell collaboration.

## Discussion

Immunosuppressive therapies used for RA can impair responses to vaccination (3). Abatacept and MTX both reduce antibody production in response to various vaccines, including the SARS-CoV-2 mRNA vaccines (15, 16). However, the cellular mechanisms by which these therapies disrupt the complex interactions required for a productive vaccine response are still poorly understood, and the impact this has on vaccine-specific T and B cell memory responses has not been characterized. Here were performed detailed phenotypic and functional characterization of vaccine-elicited T cell and B cell responses in participants with RA treated with different disease modifying therapies. We present evidence linking specific changes in T and B phenotype to reduced ability to generate anti-S antibodies after vaccination, particularly in the context of the costimulatory blockade therapy abatacept.

As expected, most individuals with RA treated with the B cell-depleting antibody rituximab had undetectable S-specific antibody responses in the serum. In addition, the S-specific CD4^+^ T cell responses were substantially reduced compared to control subjects. This lack of T cell responses in the context of rituximab treatment contrasts with prior studies of MS and B cell-depleting therapies (26, 27), and this could be due to disease-specific effects in RA vs. MS, a difference in the specific drugs used in these individuals (rituximab vs. ocrelizumab), the small size of the rituximab cohort in our study (n=5), and/or the fact that the some of the rituximab-treated subjects in our study were also on additional immunosuppressive drugs such as hydroxychloroquine or leflunomide.

In MTX-treated subjects, we observed a significantly lower magnitude of the S-specific CD4^+^ T cell response to vaccination as measured in our AIM assay. This is consistent with the known mechanism of action of MTX, which inhibits dihyrdrofolate reductase and thereby attenuates lymphocyte activation and proliferation (28). However, phenotypically and functionally, S-specific CD4^+^ T cells in the context of MTX treatment were similar and not significantly different from those observed in healthy controls. We also observed no significant changes versus healthy controls in the number of RBD-specific B cells, their phenotype, or class switching in our MTX-treated cohort, and no significant differences in either the serum S-specific antibody levels or neutralization activity. However, more highly powered studies have detected significant decreases in anti-S protein antibody levels in MTX-treated patients (29, 30), consistent with the altered T cell responses we observed in these subjects.

In contrast to the MTX-treated cohort, we found impaired SARS-CoV-2-specific CD4^+^ memory T cell, IgG^+^ memory B cell, and neutralizing antibody responses to SARS-CoV-2 vaccination in abatacept-treated RA participants (31, 32). Although the magnitude of the S-specific CD4^+^ T cell response was not significantly different than that in control subjects, we observed a significant reduction in cells producing the key cytokines IL-2, IFNγ and IL-21. Abatacept disrupts T cell activation via blockade of CD28-mediated costimulation, and we and others have consistently shown that abatacept treatment is associated with a reduction in Tfh cells (11, 12), and with an impaired transcriptional program of T cell activation and proliferation. Consistent with this, we found that CD4^+^ T cells from the abatacept-treated cohort had reduced levels of Tfh-associated cytokines, particularly IL-21. Abatacept also disrupts Tfh-B cell interactions which rely on CD28-mediated co-stimulation (33). Tfh-produced IL-21 and CD40L from these interactions are required for B cell activation and differentiation into germinal center B cells where they undergo affinity maturation and can differentiate into plasma cells producing high-affinity antibody or memory B cells poised to rapidly produce protective antibody upon a reinfection. Germinal center Tfh can produce IL-21 which particularly supports B cell differentiation into plasma cells (34, 35). We also observed decreased production of IFNγ by S-specific T cells in abatacept subjects, which promotes IgG class switching in B cells (36). Reduced SARS-CoV-2-specific CD4^+^ T cell IL-21 and IFNγ production in these subjects, and possibly abatacept directly, likely impaired T-dependent activation of SARS-CoV-2-specific B cells leading to the reduced number of SARS-CoV-2 RBD-specific antigen-experienced B cells, proportion of RBD-specific activated B cells, proportion of RBD-specific IgG^+^ B cells, and reduced neutralizing antibody that we observed.

Neutralizing antibody titers have long been considered an important correlate of protection after vaccination against viral pathogens. Therefore, our finding of reduced anti-S antibodies in the abatacept cohort is clinically relevant for understanding immune protection to SARS-CoV-2 following vaccination in these individuals. Additionally, altered formation of memory B cells is also detrimental to immune protection, as these cells are thought to be the primary reservoir of cells responding to SARS-CoV-2 variants that effectively evade vaccine-elicited neutralizing antibody responses. In our prior analyses of abatacept-treated subjects, we found that the impact of abatacept on the abundance and transcriptional profile of Tfh was rapidly reversed after drug withdrawal (11). Therefore, when clinically manageable, cessation of abatacept treatment during the course of vaccination is likely to result in significantly improved response to the COVID-19 vaccines, and defense against severe viral infection in face of future variants.

## Methods

### Peripheral blood mononuclear cell (PBMC) and plasma collection

Venous blood from study volunteers was collected in vacutainer tubes containing spray-coated silica (to prevent red cells from sticking to the tube wall) and a polymer gel for serum separation, then spun at 1400xg for 20 min. Serum was collected, heat-inactivated at 56oC for 30 min, aliquoted and stored at -80°C. The cellular fraction was resuspended in phosphate buffered saline (PBS) and PBMC were separated from red blood cells using Ficoll extraction and frozen at -80°C before being stored in liquid nitrogen. PBMCs were thawed at 37°C and washed twice before use.

### Anti-SARS-CoV-2 ELISA

Serologic testing was performed using the FDA-authorized (via Emergency Use Authorization) anti–SARS-CoV-2 IgG ELISA kit from Euroimmun (Lubeck, Germany). All testing and analyses were performed according to the manufacturer’s protocols, with the optical density ratio (ODR) calculated using the kit calibrator. The manufacturer-provided reference range is as follows: ratio <0.8, negative; ratio 0.8 to <1.1, borderline; and ratio ≥1.1, positive. To standardize results and facilitate comparisons, ODR scores for each sample were converted to z-scores (number of SDs above the negative control mean) as follows: z-score = (test ODR – mean negative control ODR) / mean negative control SD (37). Negative control sera had been collected between 2015 and 2019 from healthy community blood donors and from patients tested in the clinical laboratory by Western blot for potential HSV infection (n = 78). Based on the negative control data, ODR z-scores were therefore calculated as (ODR – 0.26)/0.13. A conservative z score ≥3 was considered positive to minimize false-positive results.

### SARS-CoV-2 RBD protein and tetramer generation

Recombinant SARS-CoV-2 RBD (from the Wuhan-1 strain, which shares an identical spike protein to the WA-1 strain) was generated as previously described (20). For tetramer generation, RBD proteins were biotinylated with the BirA500 kit (Avidity), tetramerized with streptavidin-phycoerythrin (SA-PE) (Agilent, PJRS301-1) and stored in 50% glycerol at -20°C as previously described (38). Decoy reagents were generated by tetramerizing an irrelevant biotinylated protein with SA-PE previously conjugated to Dylight594 NHS Ester (ThermoFisher, 46413) and Dylight650 NHS Ester (ThermoFisher, 62266).

### SARS-CoV-2 spike pseudotyped lentivirus

The SARS-CoV-2(WA-1)-spike pseudotyped lentivirus was produced by transient polyethylenimine transfection of 293T cells (ATCC ACS-4500, cultured in DMEM with 10% heat-inactivated FBS, 2 mM L-glutamine, 10mM HEPES, 100 U/mL penicillin, and 100 μg/mL streptomycin in a humidified atmosphere with 5% CO2 at 37°C) with a plasmid encoding the SARS-CoV-2 (WA-1) variant spike (D614G mutation and deletion of C-termina1 21aa, BEI Resources NR-53765) and additional components as described (BEI Resources NR-52516, NR-52517, NR-52518, NR-52519). Harvested supernatants were filtered through 0.2uM filters and viral titers tested as described (39).

### Pseudovirus neutralization test (pVNT)

PVNT assays were performed as previously described (39). Briefly, heat inactivated plasma was diluted 1:10 followed by four 3-fold serial dilutions all in duplicate and mixed 1:1 with 10^6^ relative luciferase units of SARS-CoV-2(WA-1)-spike pseudotyped lentivirus in DMEM with 10% heat-inactivated FBS, 2 mM L-glutamine, 100 U/mL penicillin, and 100 μg/mL streptomycin. After 1 hour incubation at 37°C, the plasma/virus mixtures were added to 96-well poly-L-lysine-coated plates seeded with human ACE2-expressing 293T cells (BEI Resources NR-52511) 20 hours prior. Each plate contained wells with no plasma and 293T cells as a background control and a plasma sample from naïve individual (collected early 2020, negative for N and RBD-specific antibodies) as a negative control (n = 4). A monoclonal anti-RBD(WA-1) antibody served as a positive control (10μg/ml starting dilution, generated by BCR sequencing single cell sorted RBD(WA-1)-specific B cells and expressing and purifying the antibody as described (40). After incubating for 48 hours, supernatant was pipet off and replaced with Bright-Glo Luciferase Assay System luciferase (Promega, E2610) for 2 min at 25°C in the dark before transferring to black-bottom plates for measuring luminescence for 1s per well on a Centro LB 960 Microplate Luminometer (Berthold Technologies). Percent neutralization was calculated as (1 – ((sample/293T-ACE2+virus RLU) – (293T+virus RLU))/((293T-ACE2+virus RLU) – (293T+virus RLU)) x 100.

### Immunophenotyping RBD-specific B cells

PBMCs were thawed at 37°C and washed twice before first staining with decoy tetramer and then with RBD tetramer prior to incubation with anti-PE magnetic beads and magnetic bead enrichment (Miltenyi Biotec, 130-048-801) as previously described (38). Cells in the positive fraction were stained with surface antibodies for B cell phenotypes (antibodies listed in Supplemental Table 1).

### Peptide Pools

SARS-CoV-2 15-mer peptides, 1mg each (BEI Resources), were provided lyophilized and stored at -80C. Peptides were selected for reactivity against a broad range of class I and class II HLA sub-types for targeted coverage of T cell epitopes as described (5, 41). Before use, peptides were warmed to room temperature for 1 hour then reconstituted in DMSO to a concentration of 10mg/mL. Individual peptides were combined in equal ratios to create Membrane/Nucleocapsid (182 ug/mL each, 55 peptides) or Spike (200ug/mL each, 49 peptides) pools, maintaining a total peptide concentration of 10mg/mL.

### T cell activation induced marker assay

For surface phenotyping 10×10^6^ PBMC per sample were divided into four 5mL polystyrene tubes and cells were pelleted at 250 x g for 5 minutes. Pellets were resuspended at 5×10^6^/mL in one of the following treatment conditions: DMSO (Sigma-Aldrich, >99.5% cell culture grade), 1μg/mL CEFX Ultra SuperStim Pool (JPT, PM-CEFX-2), or 5μg/mL SARS-CoV-2 M/N or S peptide pools. Stimulation was performed for 18 hours in ImmunoCult-XF T cell Expansion Medium (StemCell Technologies). After stimulation, cells were stained with surface antibodies for T cell activation and phenotype (antibodies listed in Supplemental Table 1).

### Intracellular cytokine assay

For intracellular cytokine assessment PBMC (3×10^6^/ml) were stimulated using either 10μg/mL SARS-CoV-2 S protein peptide pool, 50 ng/mL phorbol 12-myristate 13-acetate (Sigma-Aldrich) and 1 mg/mL Ionomycin (Sigma-Aldrich), or an equivalent volume of DMSO (Sigma-Aldrich, >99.5% cell culture grade) for 6 hours. This culture occurred in RPMI media supplemented with FCS, Penicillin/Streptomycin, sodium pyruvate, and beta-mercaptoethanol. The culture also contained anti-human CD40 antagonist mAb (Miltenyi, clone HB10) to improve resolution of CD154+ cells. 1.8uL Monensin (Becton Dickinson) was added for the final 4 hours of culture. Permeabilization and fixation was performed using Cytofix/Cytoperm (Becton Dickinson), and cells were stained with intracellular cytokine antibodies (antibodies listed in Supplemental Table 1).

### Flow cytometry

Data were acquired on a five-laser Cytek Aurora (T cell surface phenotyping and T cell intracellular cytokine analysis) or BD FACS Symphony A3 or A5 (B cell surface phenotyping). Control PBMCs or UltraComp eBeads (ThermoFisher) were used for compensation. Up to 10^7^ live PBMC were acquired per sample for T cells and all enriched PBMCs were acquired for B cells. Data were analyzed using SpectroFlow (Cytek Biosciences) and FlowJo10 (Becton Dickinson) software.

## Statistics

Statistics are described in figure legends and were determined using Prism (Graphpad). All measurements within a group are from distinct samples. Statistical significance of all pairwise comparisons was assessed by Kruskal-Wallis one-way analysis of variance with Dunn’s post-hoc test for multiple comparisons. For correlations, r-squared values are shown to indicate goodness-of-fit for linear regression and p-values are shown to indicate FDR probability of a non-zero slope. Raw p-values are displayed and the adjusted p-value significance cutoff calculated from the Benjamini-Hochberg multiple testing correction with FDR=0.05 for each figure is listed in the corresponding legend.

## Study approval

All samples were obtained upon written informed consent at the Benaroya Research Institute, part of Virginia Mason Franciscan Health in Seattle, WA, USA. All studies were approved by the Institutional Review Board of the Benaroya Research Institute (Seattle, WA).

## Data Availability

All data produced in the present study are available upon reasonable request to the authors

## Figure legends

Figure 1. Abatacept and rituximab reduce SARS-CoV-2-specific antibody levels after vaccination

(A) Study schematic and table of participant information. (B) Normalized anti-S antibody levels as measured by ELISA. (C) Pseudovirus neutralization of subjects’ sera, as area-under-curve (AUC) across serum dilutions, with historical/naive (HN) control. (D) Subject age graphed against anti-S antibody levels. (E) Time between each subject’s second vaccine dose and blood draw for the study graphed against anti-S antibody levels. Linear regression shown with r-squared values and p values testing probability of a non-zero slope. Statistics determined by Kruskal-Wallis test with post-hoc Dunn’s multiple comparison test. * p<.05, ** p<.01, *** p<.001

Figure 2. Abatacept treatment reduces activation and class-switching in RBD-specific memory B cells after vaccination

(A) Representative gating on live CD3^-^CD14^-^CD16^-^CD19^+^CD20^+^ B cells (left) and number (right) of SARS-CoV-2 RBD specific B cells (RBD tetramer^+^ Decoy tetramer^−^) from PBMCs from control (white), methotrexate (MTX, blue) and abatacept (red) treated individuals. (B) Representative gating on RBD-specific CD38^lo^ non-plasmablast B cells for naïve B cells (CD21^+^CD27^−^), classical MBCs (CD21^+^CD27^+^), activated MBCs (CD21^−^CD27^+^) and double-negative (DN) activated MBCs (CD21^−^CD27^−^). (C) Proportion of RBD-specific B cells that are each phenotype from individuals in the indicated treatment group. (D) Representative gating on RBD-specific CD38^lo^ non-plasmablast B cells for isotypes IgD, IgM, IgG and IgA. (E) Proportion of RBD-specific B cells expressing the isotypes indicated in the groups indicated. Data combined from four individual experiments. Statistics determined by Kruskal-Wallis test with post-hoc Dunn’s multiple comparison test. All statistically significant comparisons (p<.05) are shown. * p<.05, ** p<.01, *** p<.001

Figure 3. MTX and abatacept reduce S-specific memory T cell responses after vaccination

(A) Representative gating of CD3^+^CD45RA^-^CD4^+^ T cells for AIM+ (CD69^+^CD137^+^) within indicated stimulation conditions. (B) Quantification of AIM expression by subject groups as percentage of CD3^+^CD45RA^-^CD4^+^ cells. (C) Representative gating of central memory (CD45RA^-^CD27^+^), effector memory (CD45RA^-^CD27^-^), and Temra (CD45RA^+^CD27^+^) within non-naïve and AIM+ T cells. (D) Quantification of CD4 memory subsets within spike-stimulated AIM+ cells. (E) Representative gating of CXCR5^+^ (containing the Tfh population), Th1 (CXCR3^+^CCR6^-^), Th17 (CXCR3^-^CCR6^+^), Th1/17 (CXCR3^+^CCR6^+^), and Th2 (CXCR3^-^CCR6^-^CCR4^+^) cells. (F) Pie charts showing percentage of spike-stimulated AIM+ CD4 T cells falling into each Th subset. Statistics determined by Kruskal-Wallis test with post-hoc Dunn’s multiple comparison test. All statistically significant comparisons (p<.05) between treatment groups are shown. * p<.05, ** p<.01, *** p<.001

Figure 4. Abatacept-treated subjects have reduced Tfh-associated cytokine production by S-specific memory T cells after vaccination

(A) Representative gating of AIM+ (CD69^+^CD40L^+^) T cells for ICS assay co-culture. (B) Representative gating of IL-2, IL-21, IL-10, IFNγ, IL-4, IL-13, and IL-17A expression within AIM+ CD4 T cells. (C) Quantification of the expression of each cytokine by percentage of AIM+ CD4s. (D) Co-expression of IL-2, IL-21, and IFNγ in each indicated combination. (E) Anti-S antibody level graphed percent of spike-activated AIM+ CD4s expressing IL-21. (F) Anti-S antibody level graphed percent of spike-activated AIM+ CD4s expressing IFNγ. Linear regression shown with r-squared values and p values testing probability of a non-zero slope. Statistics determined by Kruskal-Wallis test with post-hoc Dunn’s multiple comparison test. All statistically significant comparisons (p<.05) are shown. * p<.05, ** p<.01, *** p<.001

Supplemental Figure 1.

(A) Percent neutralization of S-pseudotyped lentivirus infection of ACE2-expressing cells across all serum dilutions tested, with anti-RBD monoclonal antibody (mAb) as positive control and historical/naive (HN) as negative control. (B) Anti-S antibody levels graphed against pseudovirus neutralization. (C) Time between each subject’s second vaccine dose and blood draw for the study graphed against pseudovirus neutralization. (D) Age graphed against antibody Z score for abatacept subjects, split by individuals on mono vs combination therapy. All linear regression shown with r-squared values and p values testing probability of a non-zero slope.

Supplemental Figure 2.

(A) Gating strategy for identifying and phenotyping RBD-specific B cells from PBMCs. (B,C) Correlation between time since second vaccine dose and percent activated (CD21^-^CD27^+^) MBCs for control (B) and RA (C) groups. (D) Percent of atypical MBCs (CD21^-^CD27^-^CD11c^+^) of RBD-specific B cells. (E) Number of antigen-experienced (Ag-exp., CD21^+^CD27^+^ or CD21^-^ CD27^+/-^) RBD-specific B cells and (F) correlation with normalized quantity of S-specific antibody. Linear regression lines for abatacept and MTX groups. (G) Percent of RBD-specific antigen-experienced B cells expressing each isotype indicated. All linear regression shown with r-squared values and p values testing probability of a non-zero slope. Statistics determined by Kruskal-Wallis test with post-hoc Dunn’s multiple comparison test. All statistically significant comparisons (p<.05) are shown. * p<.05, ** p<.01, *** p<.001

Supplemental Figure 3.

(A) Percent AIM+ data from T cell stimulation assay shown for the membrane/nucleocapsid control condition. (B) Percent AIM+ data from T cell stimulation assay shown for the RA cohort on rituximab. Statistics determined by Mann-Whitney test. *** p<.001

Supplemental Figure 4.

Pie charts showing percentage of AIM+ CD4 T cells falling into each Th subset shown for every individual donor, with the number of AIM+ CD4s in each donor indicated.

Supplemental Figure 5.

Percentage of AIM+ (CD69^+^CD40L^+^) non-naïve CD4 T cells after DMSO or Spike stimulation in ICS assay co-culture. Kruskal-Wallis test with post-hoc Dunn’s multiple comparison test showed no significant (p<.05) differences.

Supplemental Table 1

List of antibodies (marker/fluorophore) used in each flow cytometry panel, with antibody clone and supplying company indicated.

