## Supplemental Table and Figures for "Diminished responses to mRNA-based SARS-CoV-2 vaccines in individuals with rheumatoid arthritis on immune modifying therapies"

### Figure S1

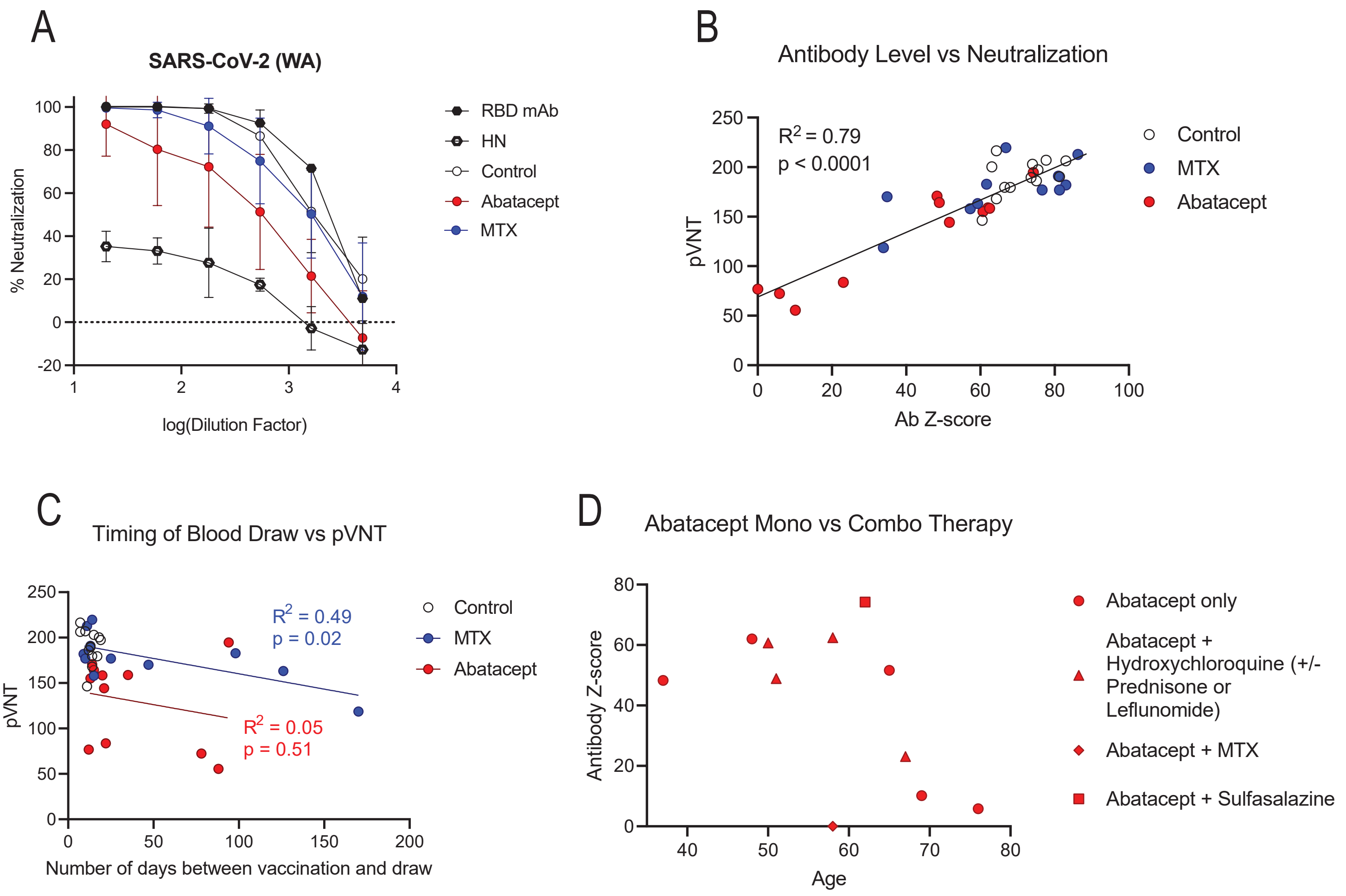

**Supplemental Figure 1.**
**(A)** Percent neutralization of S-pseudotyped lentivirus infection of ACE2-expressing cells across all serum dilutions tested, with anti-RBD monoclonal antibody (mAb) as positive control and historical/naive (HN) as negative control. **(B)** Anti-S antibody levels graphed against pseudovirus neutralization. **(C)** Time between each subject's second vaccine dose and blood draw for the study graphed against pseudovirus neutralization. **(D)** Age graphed against antibody Z score for abatacept subjects, split by individuals on mono vs combination therapy. *All linear regression shown with r-squared values and p values testing probability of a non-zero slope.*

### Figure S2

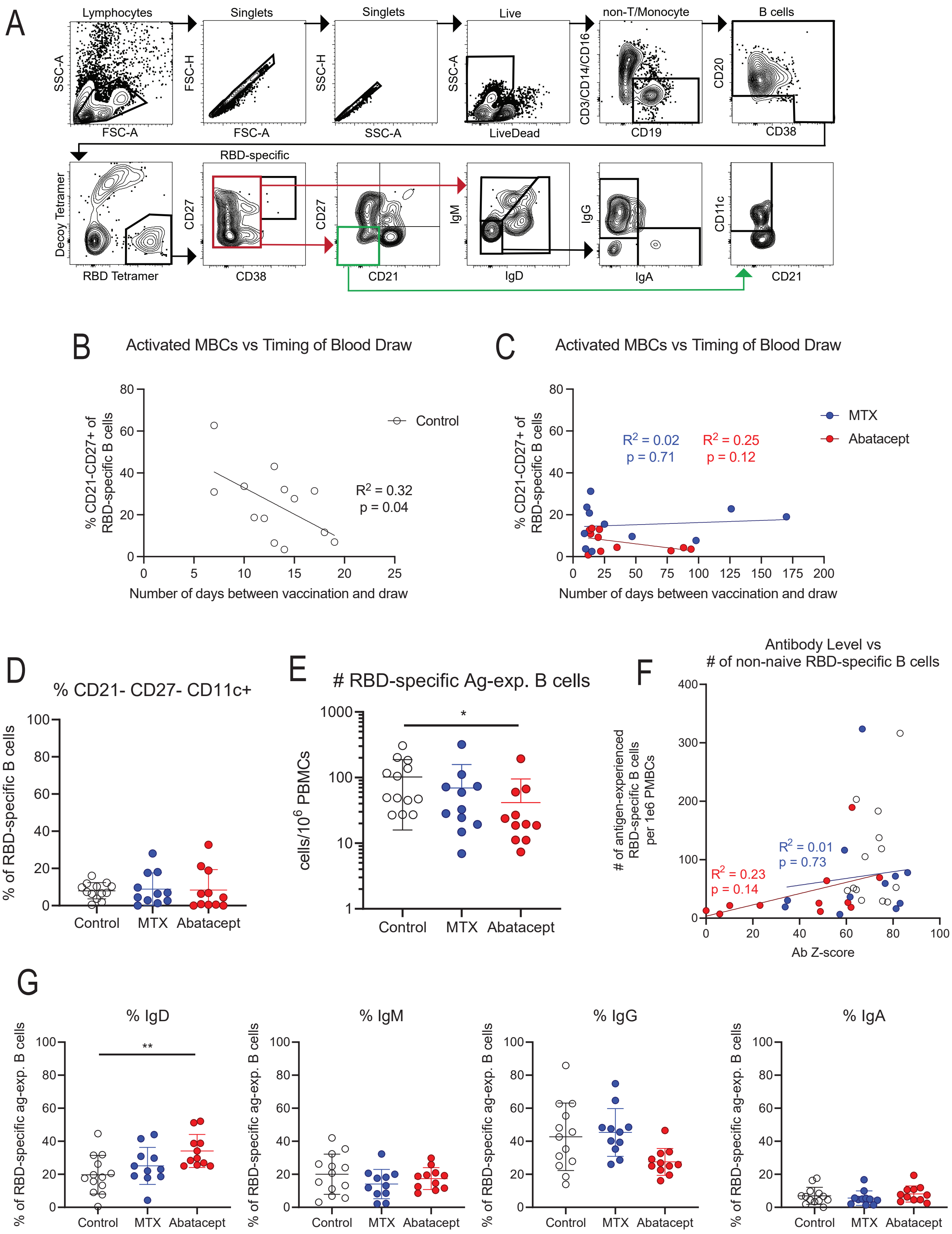

**Supplemental Figure 2.**
**(A)** Gating strategy for identifying and phenotyping RBD-specific B cells from PBMCs. **(B,C)** Correlation between time since second vaccine dose and percent activated ($CD21^-CD27^+$) MBCs for control (B) and RA (C) groups. **(D)** Percent of atypical MBCs ($CD21^-CD27^-CD11c^+$) of RBD-specific B cells. **(E)** Number of antigen-experienced (Ag-exp., $CD21^+CD27^+$ or $CD21^-CD27^{+/-}$) RBD-specific B cells and **(F)** correlation with normalized quantity of S-specific antibody. Linear regression lines for abatacept and MTX groups. **(G)** Percent of RBD-specific antigen-experienced B cells expressing each isotype indicated. *All linear regression shown with r-squared values and p values testing probability of a non-zero slope. Statistics determined by Kruskal-Wallis test with post-hoc Dunn's multiple comparison test. All statistically significant comparisons (p<.05) are shown. * p<.05, ** p<.01, *** p<.001*

### Figure S3

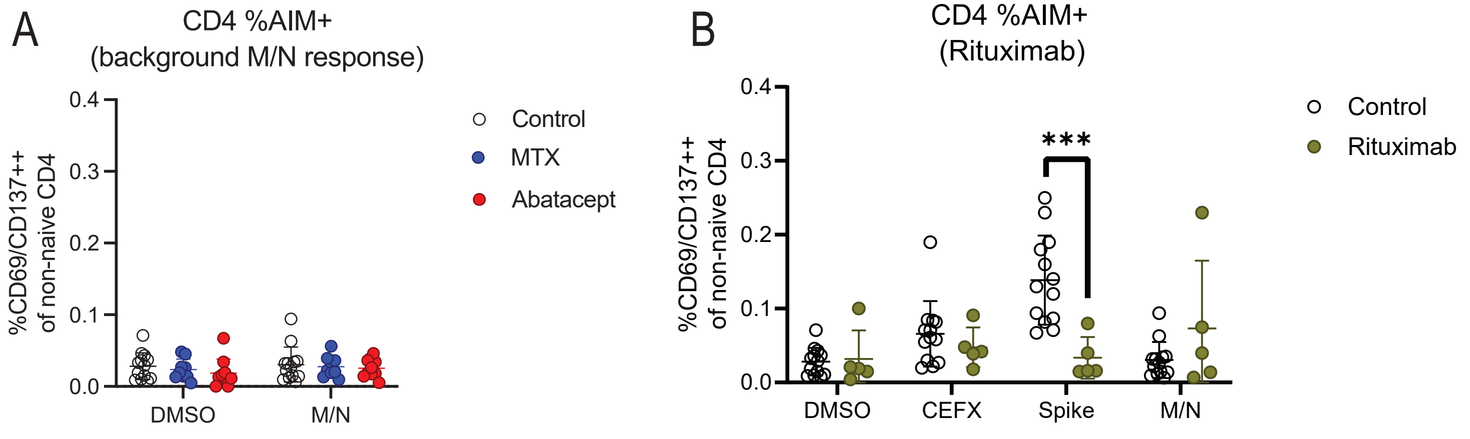

**Supplemental Figure 3.**
**(A)** Percent AIM+ data from T cell stimulation assay shown for the membrane/nucleocapsid control condition.
**(B)** Percent AIM+ data from T cell stimulation assay shown for the RA cohort on rituximab. *Statistics determined by Mann-Whitney test. *** p<.001*

Figure S4

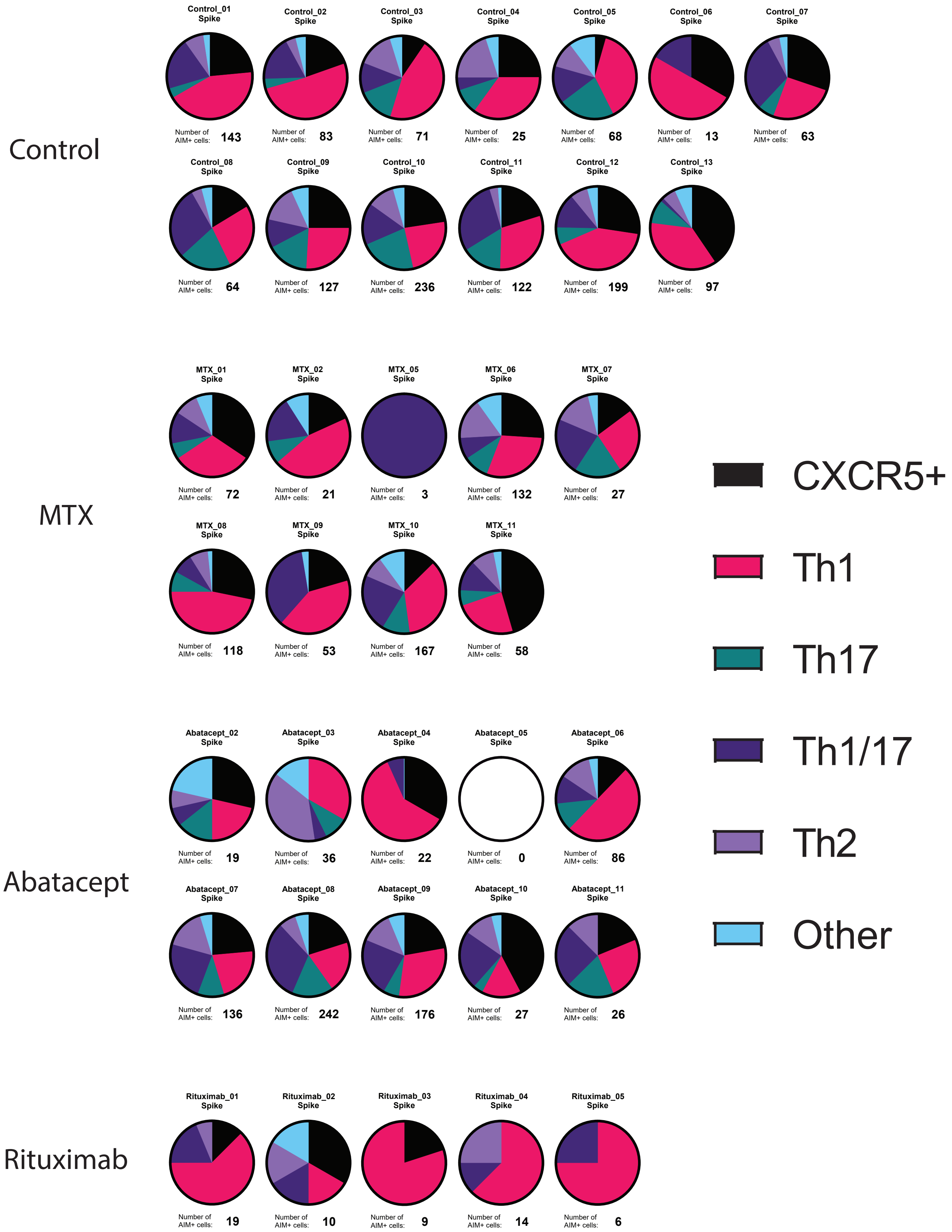

**Supplemental Figure 4.**
Pie charts showing percentage of AIM+ CD4 T cells falling into each Th subset shown for every individual donor, with the number of AIM+ CD4s in each donor indicated.

### Figure S5

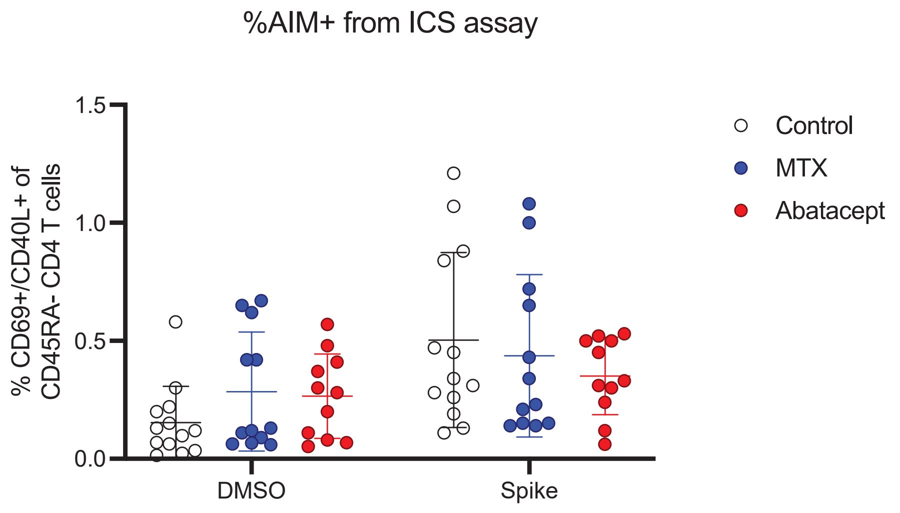

**Supplemental Figure 5.**
Percentage of AIM+ ($CD69^{+}CD40L^{+}$) non-naïve CD4 T cells after DMSO or Spike stimulation in ICS assay co-culture. *Kruskal-Wallis test with post-hoc Dunn's multiple comparison test showed no significant ($p<.05$) differences.*

### Table S1

| Panel | Antibody | Clone | Supplier |
|---|---|---|---|
| B cell panel | Live/Dead fixable blue | | Thermo Scientific |
| | CD3 :: PerCP/Cy5.5 | HIT3a | BioLegend |
| | CD14 :: PerCP/Cy5.5 | M5E2 | BioLegend |
| | CD16 :: PerCP/Cy5.5 | 3G8 | BioLegend |
| | CD19 :: BUV496 | SJ25C1 | BD Biosciences |
| | CD20 :: BV711 | 2H7 | BioLegend |
| | CD38 :: Alexa700 | 90 | ThermoFisher |
| | CD21 :: SB600 | HB5 | ThermoFisher |
| | CD27 :: BV421 | M-T271 | BioLegend |
| | IgD :: BUV395 | IA6-2 | BD Biosciences |
| | IgM :: BV510 | MHM-88 | BioLegend |
| | IgG :: BV786 | G18-145 | BD Biosciences |
| | IgA :: PE/Vio770 | IS11-8E10 | Miltenyi Biotec |
| | CD11c :: PE/Dazz594 | 3.9 | BioLegend |
| T cell AIM panel | CD45 :: BUV395 | HI30 | BD Biosciences |
| | CD45 :: BUV496 | HI30 | BD Biosciences |
| | CD45 :: efluor450 | HI30 | ThermoFisher |
| | CD45 :: Alexa532 | HI30 | ThermoFisher |
| | CD3 :: BUV615 | UCHT1 | BD Biosciences |
| | HLA-DR :: BUV661 | G46-6 | BD Biosciences |
| | CD45RA :: BUV737 | HI100 | BD Biosciences |
| | CD26 :: BUV805 | M-A261 | Thermo Scientific |
| | CXCR3 :: BV421 | 1C6 | BD Biosciences |
| | CD8a :: BV480 | RPA-T8 | BD Biosciences |
| | CCR7 :: BV605 | G043H7 | BioLegend |
| | CCR6 :: BV650 | G043G3 | BioLegend |
| | CD27 :: BV711 | M-T271 | BioLegend |
| | CD137 :: BV750 | 4B4-1 | BioLegend |
| | CD57 :: BV785 | QA17A04 | BioLegend |
| | CXCR5 :: BB515 | RF8B2 | BD Biosciences |
| | CD134 :: PerCP/Cy5.5 | BerACT35 | BioLegend |
| | PDL1 :: PE | 29E.2A3 | BioLegend |
| | CCR4 :: PE/Dazz594 | L291H4 | BioLegend |
| | CD25 :: PE/Cy5 | BC96 | BioLegend |
| | CD127 :: PE/Cy7 | hIL7Rm21 | BioLegend |
| | ICOS :: APC | C398.4a | BioLegend |
| | CD4 :: Spark685 NIR | SK3 | BioLegend |
| | CD69 :: APC/R700 | FN50 | BD Biosciences |
| | Live/Dead Zombie NIR | | BioLegend |
| | gdTCR :: APC/Fire750 | B1 | BioLegend |
| | CD19 :: APC/Fire810 | HIB19 | BioLegend |
| T cell ICS panel | CD69 :: BUV395 | FN50 | BD Biosciences |
| | Live/Dead fixable blue | | eBioscience |
| | IL-13 :: BV421 | JES10-5E2 | BioLegend |
| | CD3 :: efluor450 | OKT3 | eBioscience |
| | CD107a :: BV510 | H4A3 | BioLegend |
| | IL-17A :: BV570 | BL168 | BioLegend |
| | CD40L :: Biotin | hCD40L-M91 | BD Biosciences |
| | Streptavidin :: BV605 | 563260 | BD Biosciences |
| | CD25 :: BV650 | M-A251 | BD Biosciences |
| | CD19 :: BV711 | SJ25C1 | BD Biosciences |
| | CD16 :: BV711 | 3G8 | BD Biosciences |
| | CD14 :: BV711 | MOP9 | BD Biosciences |
| | CD45RA :: BV711 | HI100 | BD Biosciences |
| | IL-2 :: BV785 | MQ1-17H12 | BioLegend |
| | CD127 :: Alexa488 | AO19D5 | BioLegend |
| | IL-21 :: PE | 3A3-N2 | eBioscience |
| | IL-10 :: PE/Dazz594 | JES3-9D7 | BioLegend |
| | CD8 :: PE/Cy5 | RPA-T8 | BD Biosciences |
| | IL-4 :: PE/Cy7 | MP4-25D2 | BioLegend |
| | CXCR5 :: Alexa647 | J252D4 | BioLegend |
| | CD4 :: Alexa700 | RPA-T4 | BD Biosciences |
| | IFNγ :: APC/efluor780 | 4S-B3 | eBioscience |

**Supplemental Table 1.**
List of antibodies (marker/fluorophore) used in each flow cytometry panel, with antibody clone and supplying company indicated.
